# Transplacental transfer efficiency and longitudinal dynamics of antibodies against RSV in Chinese children from birth to 8 years: a paired mother–neonate cohort study

**DOI:** 10.1101/2025.10.15.25338064

**Authors:** Meng Xu, Qianli Wang, Yan Wang, Xiaofeng Tong, Lan Yi, Jiqun Lin, Junyi Zou, Sihong Zhao, Juan Yang, Helen Y. Chu, Hongjie Yu

**Affiliations:** Shanghai Institute of Infectious Disease and Biosecurity, School of Public Health, Fudan University, Shanghai, China; Anhua County Center for Disease Control and Prevention, Yiyang, China; Department of Medicine, Division of Allergy and Infectious Diseases, University of Washington, Seattle, Washington, USA; Fudan University, Key Laboratory of Public Health Safety, Ministry of Education, Shanghai, China; Department of Infectious Diseases, Huashan Hospital, Fudan University, Shanghai, China

**Author notes:** Corresponding author: Hongjie Yu, Shanghai Institute of Infectious Disease and Biosecurity, School of Public Health, Fudan University, Shanghai, China. Meng Xu, Qianli Wang, and Yan Wang contributed equally to this work.

## Abstract

Maternal antibodies protect infants from severe respiratory syncytial virus (RSV) infection in early life, but this immunity is short-lived, resulting in increasing infection risk over time. With the approval of the first maternal vaccine and half-life extended monoclonal antibodies (mAb) for infants, understanding transplacental transfer efficiency and RSV antibody dynamics is essential for optimizing immunization strategies. A longitudinal paired mother-neonate cohort study was conducted in southern China from 2013 to 2021, with serum samples collected from mothers at delivery and from neonates at birth, and subsequently at 2, 4, 6, 12, 24, 36 months and 5-8 years of age. All samples were tested for RSV pre-F IgG antibodies using an enzyme-linked immunosorbent assay (ELISA). A total of 695 mother-neonate pairs were enrolled in this study. A strong positive correlation (ρ = 0.87; *P*<0.001) was observed between maternal antibody titers at delivery (geometric mean titer [GMT]: 4.74; 95% confidence interval [95% CI]: 4.72-4.76) and neonatal antibody titers at birth (GMT: 4.90; 95% CI: 4.88-4.92). The mean transplacental transfer ratio was estimated to be 1.44 (95% CI: 1.40-1.47). Maternal antibody titers and gestational age at birth were significantly associated with neonatal antibody levels. A ten-fold increase in maternal antibody titers was associated with a 5.46-fold increase in neonatal antibody titers (95% CI: 4.89 - 6.08; *P*<0.001). Compared with full-term birth (gestational age 37-42 weeks), pre-term birth (gestational age < 37 weeks) was associated with a 0.17-fold decrease in neonatal antibody titers (95% CI: 0.07-0.26; *P*=0.003). We observed a rapid decline of maternally acquired antibodies in neonates after birth, with a 34.3 days half-life (95% CI: 33.9-34.8). Neonatal antibody titers declined to their lowest point (3.16, 95% CI: 3.08 - 3.24) at a mean age of 10 months; this was followed by a gradual increase due to natural infection, rebounding to levels comparable to those at birth by 5-8 years of age (GMT: 4.91, 95% CI: 4.86-4.95). These findings emphasize the urgent need for targeted immunization strategies to bridge the RSV vulnerability gap during early childhood.

## Introduction

Respiratory syncytial virus (RSV) is the leading cause of lower respiratory tract infections (LRTIs) in infants and young children, globally,^1,2^ with 33 million annual cases in children under 5 years, predominantly in low- and middle-income countries.^3-5^ Previous studies reported the highest incidence of RSV infections in children under 6 months of age, particularly those younger than 3 months.^5^ Lifelong reinfections occur due to lack of protective immunity induced by natural infection.^6^

Despite limited treatment options, significant progress has recently been made in RSV preventive strategies, with a focus on the prefusion F (pre-F) protein as a key antigenic target. Currently, three licensed RSV preventive strategies have been implemented for protecting infants and young children. The first maternal RSVpreF vaccine, Abrysvo, was approved in 2023 for use in the US at 32-36 weeks of gestation and in Europe at 24-36 weeks, aiming to prevent RSV-associated LRTI in infants under 6 months.^7,8^ Additionally, two long-acting monoclonal antibodies (mAbs), nirsevimab and clesrovimab, now serve as passive immunization strategies for pediatric populations.^9,10^ Both approaches have demonstrated high effectiveness in preventing RSV LRTI in infants in recent studies.^11-13^

Maternally acquired antibodies are essential for protecting infants against severe RSV infection in early life. The maternal RSV vaccine relies on efficient transplacental transfer to pass antibodies to their newborns. Previous studies have reported considerable variability in transfer efficiency of neutralizing antibodies and total RSV IgG antibodies.^14-17^ However, our understanding of this key parameter for the maternally - derived RSV pre-F antibodies in the general population are limited. Moreover, transfer efficiency is influenced by multiple factors, such as maternal antibody levels and gestational age at delivery, highlighting the need to quantify these determinants to guide maternal immunization strategies.^14,15^

After birth, the waning of maternally acquired immunity accompanies a gradually increasing risk of natural RSV infection in infants. Although studies have shown that administration of RSV mAbs can effectively reduce the risk of severe infection, more robust data on antibody kinetics are needed to optimize passive immunization strategies, particularly in China - a country with one of the highest RSV burdens globally - where nirsevimab has been approved but is not yet widely implemented.^18^ Previous studies have characterized population-level RSV immunity through cross-sectional serological surveys,^21^ or described early-life dynamics of maternal antibodies in mother-neonate cohorts.^14,19,20^ However, long-term longitudinal antibody dynamics in the general pediatric population have not been well characterized.

Crucial evidence is still needed to guide and optimize RSV preventive strategies in infants and children. In this study, we measured RSV pre-F IgG antibody titers in serum samples from a mother-neonate cohort in Anhua County, Hunan province, southern China, from 2013 to 2021. We evaluated the transplacental transfer efficiency of maternal RSV pre-F IgG antibodies and quantified the impact of associated factors. We also characterized the longitudinal dynamics of antibody levels from birth through 8 years of age.

## Results

### Study participants

A total of 695 mother-neonate pairs, comprising 687 mothers and 695 neonates (including eight pairs of twins), were included in this study from an original cohort of 1,066 pairs. Blood samples were collected from mothers at delivery and cord blood from neonates at birth, and subsequently at 2, 4, 6, 12, 24, 36 months and once when children were 5-8 years of age. Details of the cohort design have been published elsewhere^21^. From the original 1,066 pairs, we first included all mother-neonate pairs with either follow-up data at 5-8 years of age (239 pairs) or preterm neonates (27 pairs) to ensure sufficient sample size in these subgroups, then randomly sampled from the remaining pairs for inclusion. The follow-up rates for the selected participants ranged from 56.8% to 88.9% across the first six follow-up visits between 2013 and 2018, with a subsequent rate of 34.4% at the most recent follow-up in 2021 (Fig. 1 and Fig. S1). Baseline maternal and neonatal characteristics were generally comparable between selected and non-selected mother-neonate pairs (*P*>0.05), except for gestational age at birth (*P*<0.001), as all preterm infants were intentionally included in this subset (Table S1).

**Fig. 1.**
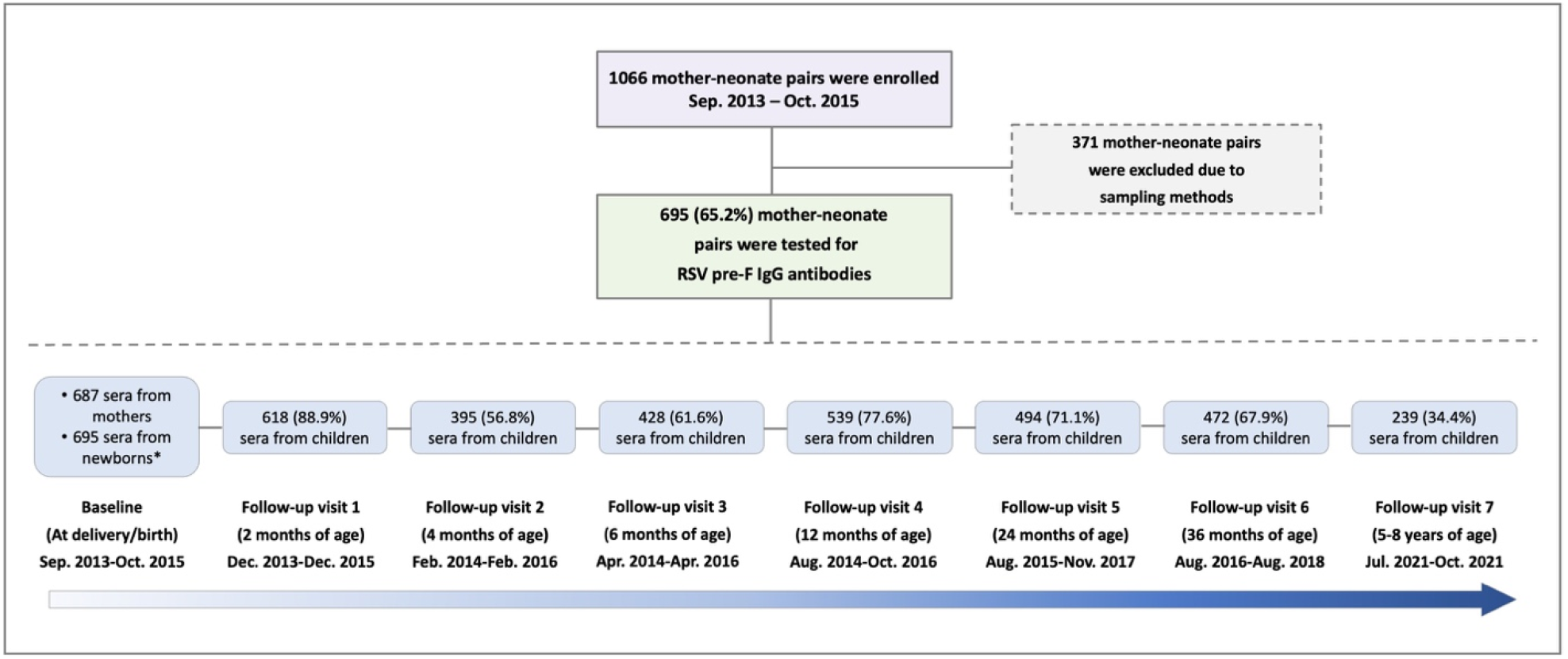
Enrollment and follow-up of study participants. * Eight pairs of twins were included in this study. The follow-up rate at each visit between 2013 and 2021 was calculated with a denominator of 695.

Among all enrolled mothers, the median age at delivery was 25 years (interquartile range [IQR]: 23–29). Among neonates, 52.8% (367/695) were male; the median gestational age at birth was 40.0 weeks (IQR: 39.3-40.7), with 3.9% (27/695) preterm births (gestational age < 37 weeks), 92.7% (644/695) full-term births (gestational age 37-42 weeks), and 3.5% (24/695) post-term birth (gestational age > 42 weeks), respectively (Table 1).

**Table 1.**
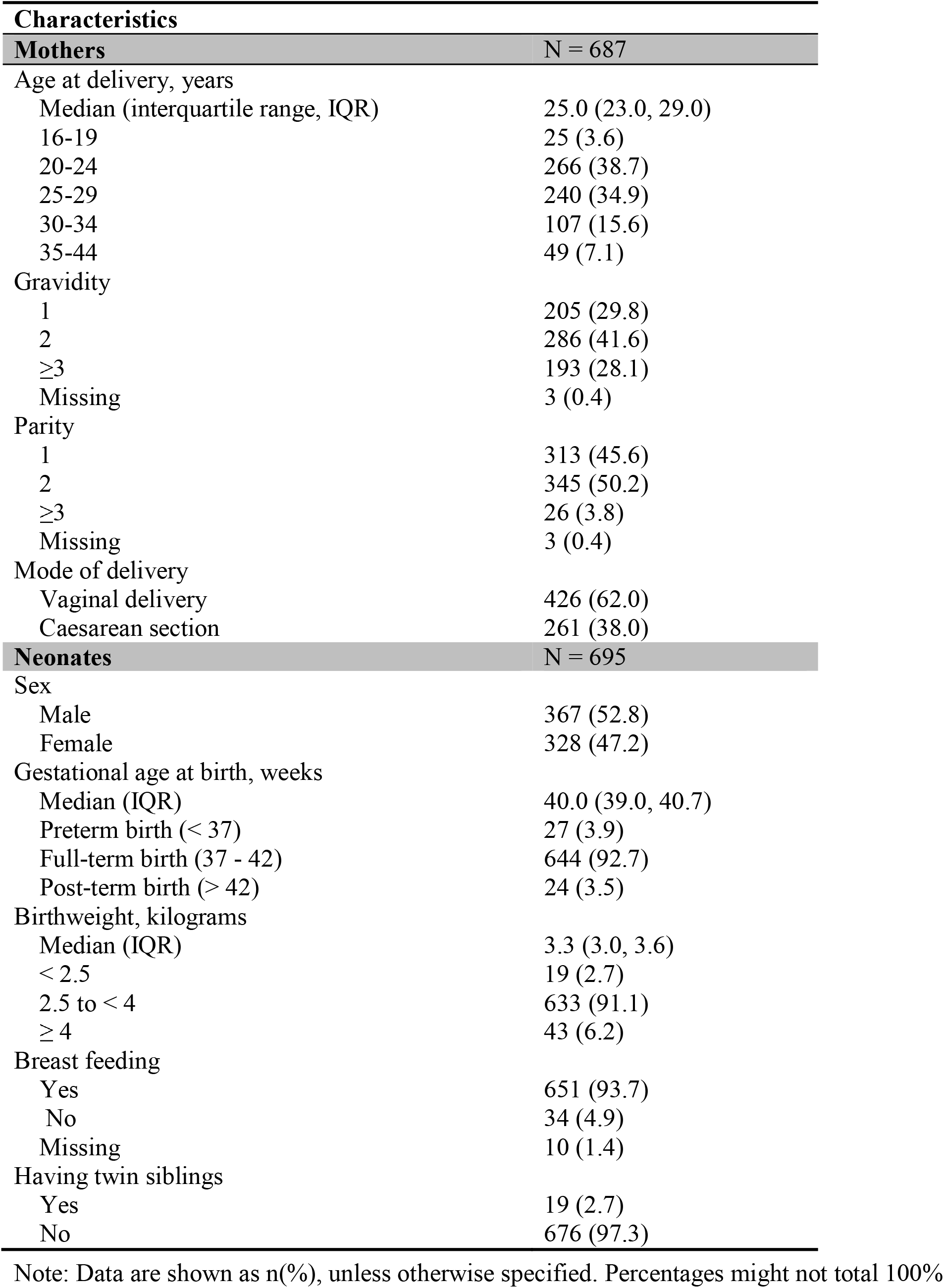
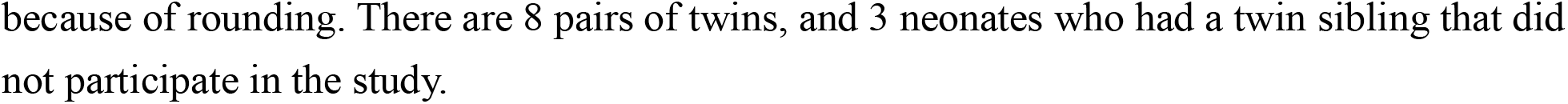
Characteristics of study participants at baseline.

| **Characteristics** |  |
| --- | --- |
| **Mothers** | **N = 687** |
| Age at delivery, years |  |
| Median (interquartile range, IQR) | 25.0 (23.0, 29.0) |
| 16-19 | 25 (3.6) |
| 20-24 | 266 (38.7) |
| 25-29 | 240 (34.9) |
| 30-34 | 107 (15.6) |
| 35-44 | 49 (7.1) |
| Gravidity |  |
| 1 | 205 (29.8) |
| 2 | 286 (41.6) |
| ≥3 | 193 (28.1) |
| Missing | 3 (0.4) |
| Parity |  |
| 1 | 313 (45.6) |
| 2 | 345 (50.2) |
| ≥3 | 26 (3.8) |
| Missing | 3 (0.4) |
| Mode of delivery |  |
| Vaginal delivery | 426 (62.0) |
| Caesarean section | 261 (38.0) |
| **Neonates** | **N = 695** |
| Sex |  |
| Male | 367 (52.8) |
| Female | 328 (47.2) |
| Gestational age at birth, weeks |  |
| Median (IQR) | 40.0 (39.0, 40.7) |
| Preterm birth (< 37) | 27 (3.9) |
| Full-term birth (37 - 42) | 644 (92.7) |
| Post-term birth (> 42) | 24 (3.5) |
| Birthweight, kilograms |  |
| Median (IQR) | 3.3 (3.0, 3.6) |
| < 2.5 | 19 (2.7) |
| 2.5 to < 4 | 633 (91.1) |
| ≥ 4 | 43 (6.2) |
| Breast feeding |  |
| Yes | 651 (93.7) |
| No | 34 (4.9) |
| Missing | 10 (1.4) |
| Having twin siblings |  |
| Yes | 19 (2.7) |
| No | 676 (97.3) |

Note: Data are shown as n(%), unless otherwise specified. Percentages might not total 100%
because of rounding. There are 8 pairs of twins, and 3 neonates who had a twin sibling that did not participate in the study.

### Transplacental transfer efficiency of maternal antibodies

We first assessed the transplacental transfer efficiency of maternal RSV pre-F IgG antibodies using paired serum samples collected from mothers at delivery and their neonates at birth. A strong positive correlation was observed between maternal and neonatal antibody levels (*ρ* = 0.87, *P* < 0.001). Notably, the geometric mean titer (GMT, log-transformed) of newborns (4.90, 95% CI: 4.88-4.92) was higher than that of the mothers (4.74, 95% CI: 4.72-4.76, Fig. 2A). Among mothers, GMTs were slightly higher in the preterm group (GMT = 4.82, 95% CI: 4.69-4.95) compared with the full-term (GMT = 4.73, 95% CI: 4.71-4.76) and post-term (GMT = 4.76, 95% CI: 4.65-4.87) groups, whereas neonatal GMTs remained similar across all gestational age groups (Fig. 2B). The transplacental transfer efficiency, defined as the geometric mean ratios of neonate-to-mother titers, was estimated to be 1.44 (95% CI: 1.40-1.47). A modest negative correlation was observed between maternal antibody titers and transplacental transfer ratios (*ρ* = −0.27, *P* < 0.001; Fig. 2C). Transfer ratios increased from 1.17 (95% CI: 1.02-1.34) in the preterm group to 1.45 (95% CI: 1.41-1.48) in the full-term group and 1.45 (95% CI: 1.18-1.79) in the post-term group.

**Fig. 2.**
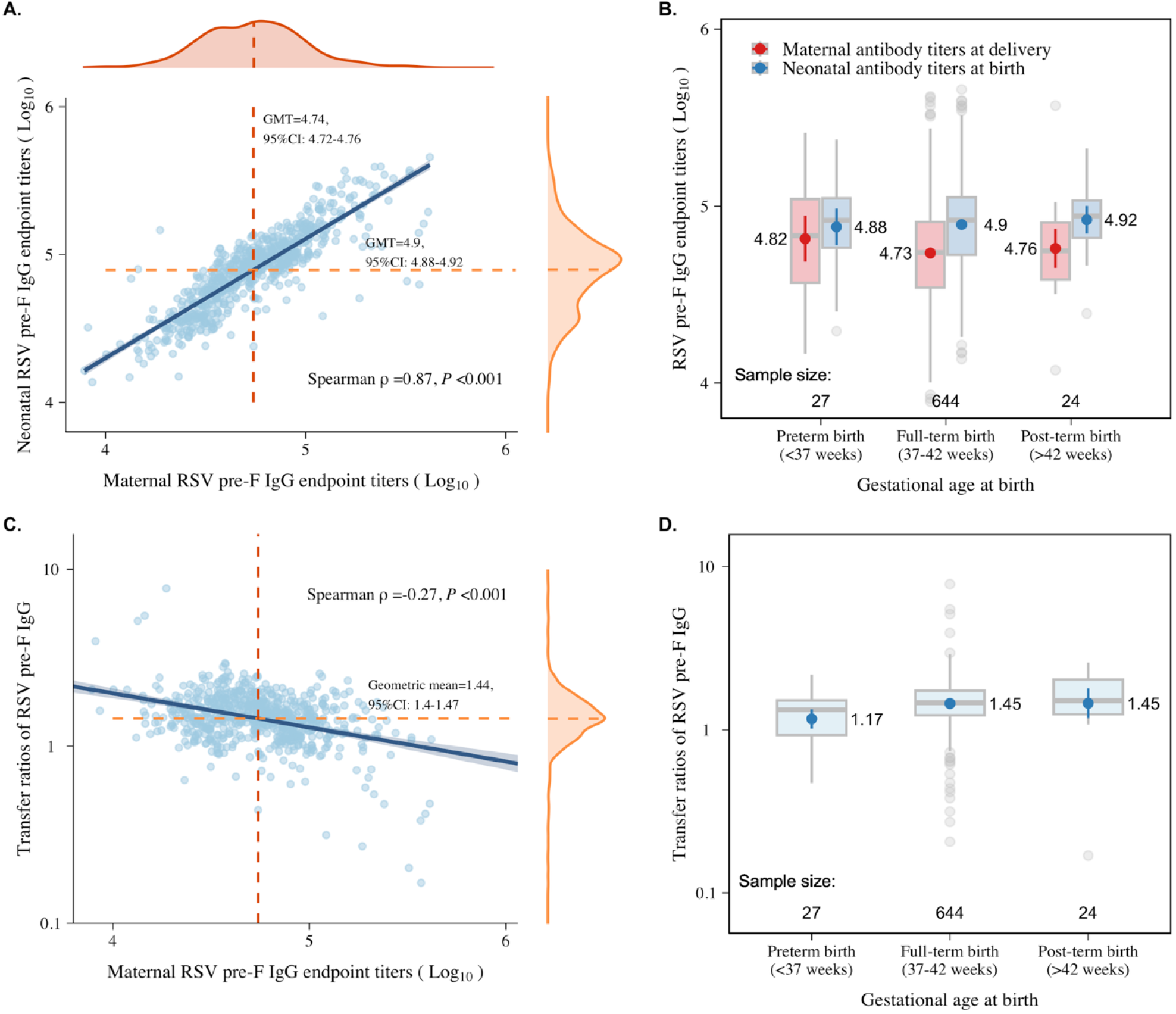
Transplacental transfer of maternal RSV pre-F IgG antibodies. (A) Correlation between maternal antibody titers at delivery and neonatal antibody titers at birth. (B) Maternal and neonatal antibody GMTs in different gestational age groups. (C) Correlation between maternal antibody titers and transplacental transfer ratios. (D) Transplacental transfer ratios of maternal antibodies in different gestational age groups.

Generalized linear models (GLMs) were then used to explore risk factors associated with neonatal antibody levels and transplacental transfer ratios. Univariate analysis suggested that maternal antibody levels potentially influenced neonatal antibody levels, while both maternal antibody levels and gestational age were potential factors influencing transfer efficiency (Tables S2 and S3). Multivariate analysis demonstrated that every ten-fold increase in maternal antibody levels was associated with a 5.46-fold increase in neonatal antibody levels, corresponding to a 0.35-fold decrease in transplacental transfer ratios (see Method section for details). Preterm birth was associated with a 0.17-fold decrease in neonatal antibody levels after accounting for maternal antibody levels (Table 2).

**Table 2.** Factors associated with neonatal RSV pre-F IgG antibodies at birth.

| Characteristics | $\beta$ (95% CI) | Fold changes<br>(95% CI) | *P* value |
| --- | --- | --- | --- |
| Maternal antibody titers | 0.81 (0.77, 0.85) | 5.46 (4.89, 6.08) | <0.001 |
| Gestational age at birth, weeks |  |  |  |
| Preterm birth (< 37) | -0.08 (-0.13, -0.03) | -0.17 (-0.26, -0.07) | 0.003 |
| Full-term birth (37 - 42) | Reference | Reference | - |
| Post-term birth (> 42) | 0.01 (-0.05, 0.06) | 0.02 (-0.11, 0.15) | 0.812 |

Note: We quantified RSV pre-F IgG antibodies using $\log_{10}$ endpoint titers. $\beta > 0$ indicates that the factor was associated with an increase in the corresponding outcomes by $10^\beta$ -1 fold; $\beta < 0$ indicates that the factor was associated with a decrease in the corresponding outcomes by $1-10^\beta$ fold.

### Dynamic pattern of maternally and naturally acquired antibodies

Next, we characterized the longitudinal dynamics of neonatal antibody levels from birth through 8 years of age. The GMTs of RSV pre-F IgG antibodies declined from 4.90 (95% CI: 4.88-4.92) at birth to 3.25 (95% CI: 3.17-3.33) by 12 months of age, then gradually increased to 4.01 (95% CI: 3.91-4.10) at 2 years and 4.45 (95% CI: 4.37-4.53) at 3 years. By 5-8 years of age, the GMT returned to 4.91 (95% CI: 4.86-4.95), a level comparable to that observed at birth (Fig. 3A). A generalized additive mixed-effects model (GAMM) was applied to fit antibody dynamics from birth to 42 months of age (Fig. 3B). A rapid decline of maternally acquired antibodies was observed during the first few months of life. Antibody levels reached their lowest point (GMT: 3.16, 95% CI: 3.08-3.24) at a mean age of 10 months, followed by a gradual increase due to natural infection.

**Fig. 3.**
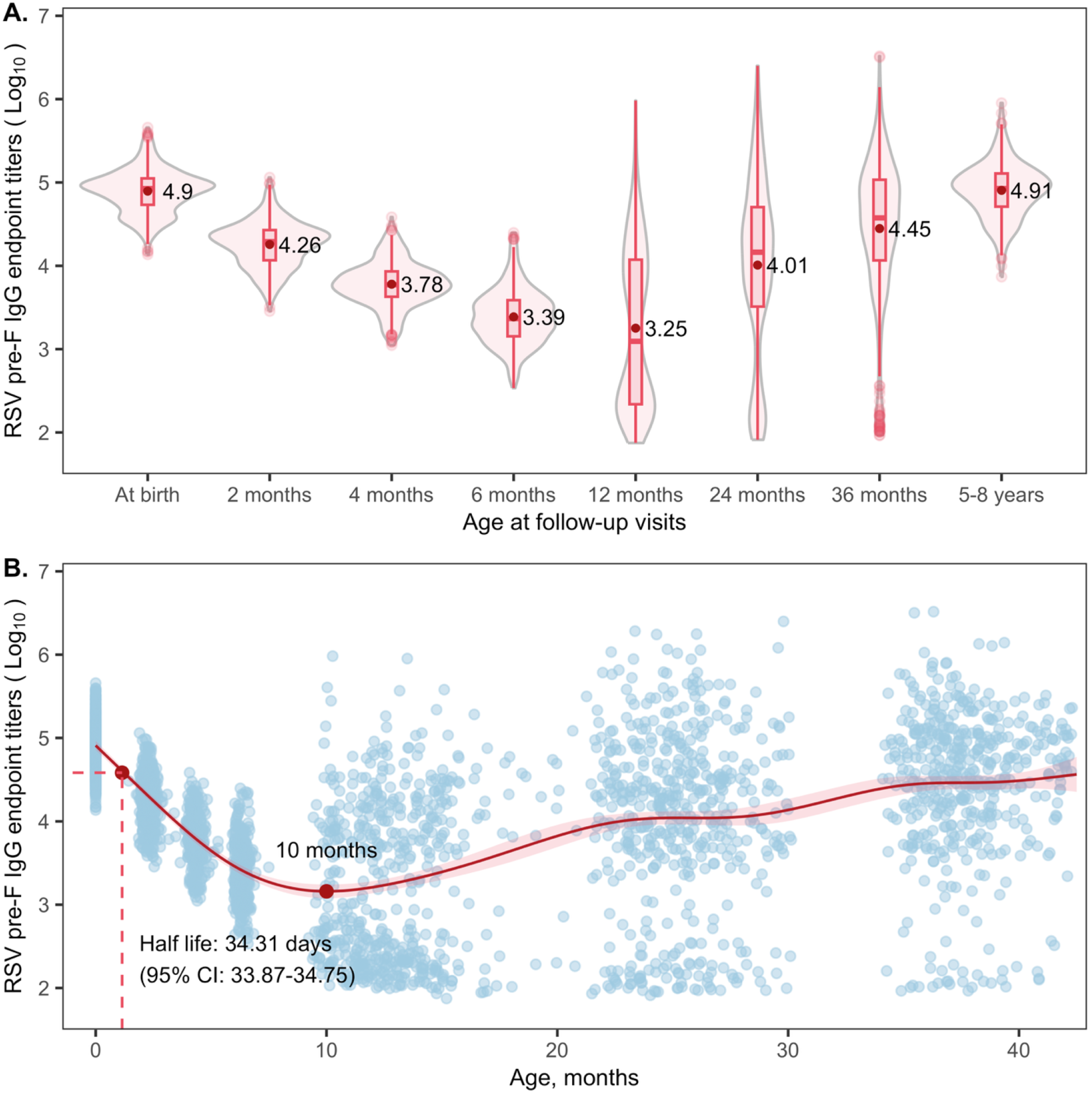
Longitudinal dynamics of RSVpre-F IgG antibody levels. (A) Age-specific GMTs in 0-8 year age groups. (B) Individual antibody titers at each visit and fitted antibody dynamics from birth through 42 months of age.

Using neonatal data collected within the first 6 months and a generalized linear mixed-effects model (GLMM), the half-life of maternal RSV pre-F IgG antibodies was estimated to be 34.3 days (95% CI: 33.9-34.8). The correlation between maternal and neonatal antibody levels declined with age: the Spearman correlation coefficients *ρ* = 0.83 at 2 months (*P* < 0.001), *ρ* = 0.73 at 4 months (*P* < 0.001), and *ρ* = 0.53 at 6 months (*P* < 0.001, Fig. S2).

Notably, most serum samples in this study were collected between 2013-2018, prior to the COVID-19 pandemic. However, the last follow-up visit (i.e., all participants aged 5-8 years) was conducted in 2021, when these participants had experienced minimal RSV exposure for nearly two years due to pandemic-related restrictions. To more thoroughly assess the potential impact of the pandemic on our findings, we randomly selected 1977 samples from children under 8 years of age from two other independent community-based studies in Anhua (see Supplementary Methods for details) and compared RSV pre-F IgG antibody titers before and after the pandemic. In children under 2 years of age, antibody levels measured in 2021 were lower than pre-pandemic levels in each age group, whereas levels in children aged 3-8 years remained comparable (Fig. S3).

### Correlation between RSV pre-F IgG and nAbs

Neutralization antibodies serve as a reliable correlate of protection (CoP) for RSV infection, and neutralization assays, which directly quantify functional antibody activity, remain the golden standard for assessing this. However, previous studies have demonstrated that the magnitude of RSV neutralizing activity is determined by pre-F IgG antibody levels^14,22,23^ Therefore, we further evaluated RSV nAb titers in a subset of 167 mother-neonate pairs, comprising 1,327 serum samples (Table. S4) and found a strong correlation between RSV pre-F IgG levels and nAb titers (*ρ* = 0.904, *P* < 0.001; Fig. S4).

## Discussion

In this study, we demonstrated efficient transplacental transfer of maternal RSV pre-F IgG antibodies. Multivariable models demonstrated that maternal antibody levels and gestational age were significantly associated with neonatal antibody levels at birth. At the study population level, antibody titers reached their lowest point at around 10 months after birth, then gradually increased through natural infection, and rebounded to levels comparable to those at birth by 5-8 years of age. These findings highlight the role of maternal RSV antibodies in early-life protection against RSV infection and advance our understanding of neonatal RSV immunity, particularly in the context of newly introduced maternal vaccines and the ongoing development of other immune interventions.

Maternally acquired antibodies are critical for protecting infants against RSV infection. In our study, RSV pre-F IgG antibodies were transferred from mothers to newborns with remarkable efficiency, with a transfer ratio of 1.44 (95% CI: 1.40-1.47), comparable to the ratio of 1.6 (95% CI: 1.47 to 1.75) observed in the placebo group of the maternal RSVPreF3 (Arexvy) vaccine trial.^24^ Compared to previous studies, the transfer ratio of RSV pre-F IgG antibodies observed here was higher than those reported for nAbs (0.60 - 1.37) and total RSV IgG antibodies (1.09 −1.22)^14,20,25-34^. Notably, the placebo group in a phase 2 clinical trial of the RSVpreF (Abrysvo) vaccine in pregnant women showed an even higher nAbs transfer ratio of 1.92 (95% CI: 1.56-2.36) ^35^, highlighting heterogeneity across studies and populations. These variations are likely attributable to methodological differences in antibody assays and variability in maternal age, gestational age, and timing of sample collection.

Our study revealed a strong correlation between maternal and neonatal pre-F IgG antibody titers at delivery (*ρ* = 0.87), consistent with prior findings for RSV total IgG titers (0.85-0.87), suggesting maternal pre-F IgG levels reliably predict neonatal antibody titers at birth.^20,36^ Multivariable analysis showed maternal antibody titers and gestational age were significantly associated with neonatal antibody levels. Higher maternal pre-F IgG levels were associated with elevated neonatal titers and a reduced transfer ratio, potentially due to placental transfer saturation of the neonatal Fcγ receptor.^37^ Furthermore, prematurity was identified as a significant factor that negatively impacted neonatal antibody titers, associated with a 0.17-fold decrease in neonatal antibody titers compared to full-term birth after adjusting for maternal antibody levels. This finding highlights the vulnerability of preterm infants, potentially increasing their susceptibility to RSV infection. However, the exact protective threshold for RSV immunity and whether it varies for preterm infants remain unknown. Despite this knowledge gap, it still emphasizes the urgent need for maternal immunization or monoclonal antibody strategies, particularly in populations with higher prematurity rates, to enhance neonatal protection against RSV. It also supports policy recommendations that preterm infants receive a dose of mAb at birth, irrespective of their maternal vaccination status.

We found that as maternal antibodies in neonates gradually waned, the levels of RSV pre-F IgG antibodies progressively increased with age, primarily driven by natural infection. This trend is consistent with findings from a cross-sectional study in a Dutch population^38^, supporting the fact that natural exposure to RSV plays a critical role in formation of infant immune response. We demonstrated a half-life of maternal RSV pre-F IgG antibodies to be 34.3 days (95% CI: 33.9-34.8), consistent with previous studies (38-42 days).^38,39^ However, previous studies reported the lowest RSV nAbs and total RSV IgG titers at 6 to 7 months ^14,20,32,40^, which differed from the findings of our study where we found this to be 10 months. This discrepancy in the timing of the lowest antibody titer may be attributed to differences in the duration and timing (season) of follow-up across studies, as well as differential exposure to RSV through the first year of life across the two cohorts. The positive correlation between maternal and neonatal antibody levels at 2, 4, and 6 months demonstrates the sustained influence of maternal antibodies on infant immunity after birth. However, as infants grow, the impact of maternal antibodies gradually wanes, underscoring the need for additional protective strategies (e.g., prophylactic mAbs) to prevent RSV infection (Fig. S2).

Notably, our results were consistent with a previous longitudinal study that evaluated the impact of reduced virus transmission during the COVID-19 pandemic on population immunity, which demonstrated no evidence of a significant decline in anti-RSV antibodies among individuals over 12 months of age^41^. This highlights age-specific differences in immune response dynamics under conditions of reduced viral circulation (Fig. S3).

In addition, our neutralization assay also revealed a robust correlation between RSV pre-F IgG and nAbs (Fig. S4), demonstrating that the longitudinal antibody dynamics described in this study are a reliable indicator of study population-level immune protection. This finding highlights the interest for further studies to validate the utility of RSV pre-F ELISA as a reliable surrogate for neutralization assays.

Our study has several limitations. First, the lack of active disease monitoring limited the reliable determination of individual infection status, thereby restricting our ability to evaluate RSV infection incidence. Testing for serum IgA may partially enhance diagnostic capacity. However, serum IgA is transiently elevated at the time of infection. Given the follow-up design of this study, it is unlikely that we would be able to use IgA rise as a surrogate of acute infection. Future studies could consider incorporating RSV IgA measurements to provide a more comprehensive analysis of RSV exposure and immunity. Second, we measured only RSV pre-F IgG antibody levels in all serum samples and RSV nAbs in a subset of samples; however, it is possible that other non-neutralizing antibody (e.g. IgA) and Fc-mediated effector functions such as antibody-dependent cellular cytotoxicity (ADCC) and antibody-dependent cellular phagocytosis (ADCP) activities may play a role in protection against disease in children. Third, we observed a significant impact of preterm birth (gestational age < 37 weeks) on the transfer efficiency of maternal pre-F IgG antibodies against RSV. However, due to the limited sample size (only 27 preterm infants), we were unable to perform more detailed subgroup analyses across different gestational age categories to understand if this affect is limited to those born at less than 37 weeks. Moreover, the absence of follow-up data between 2018 and 2021 due to the COVID-19 pandemic restricted our ability to describe antibody dynamics in children aged 36 to 60 months. Last, our results may have been influenced by factors that increase the likelihood of RSV exposure, such as daycare or preschool attendance among young children. Unfortunately, information on these specific factors was not available in our study.

In summary, this study demonstrated efficient transplacental transfer of maternal RSV pre-F IgG antibodies to neonates, but less efficiency in preterm infants. It revealed the rapid waning of maternally acquired immunity over time and demonstrates population-level RSV antibody dynamics over several years of follow up. Our findings support the importance of, and provide key evidence for the optimization of, maternal immunization strategies to protect neonates and further preventive approaches to protect young children.

## Data Availability

The original database containing confidential individual information cannot be made public. The data that support the findings of this study are available from the corresponding author upon reasonable request.

## Methods

### Study design and participants

A community-based mother-infant cohort was originally established to investigate pediatric enterovirus A71 infections in Anhua County, Hunan Province, a rural area in southern China. Details of the cohort have been described previously.^42-44^ Briefly, 1066 pregnant women and their newborns pairs were enrolled at local hospitals between September 2013 and October 2015. The neonates were followed up at the ages of 2, 4, 6, 12, 24, and 36 months from 2013 to 2018, with an additional follow-up visit conducted in July to October 2021 when they were all between 5 and 8 years old. At enrollment (time of delivery), peripartum venous blood samples were collected from mothers and cord blood samples were collected from newborns at birth. Thereafter, 2 mL of venous blood was collected from the infants at each follow-up visit. Baseline and follow-up data was collected using structured questionnaires, including demographics, gestational week of birth, delivery method (vaginal, caesarean section), and birth weight.^42-44^.

We determined that a sample size of 598 mother-neonate pairs would enable the estimation of a 46.5% annual risk of RSV infection within a margin of ± 4%, at a significance level of 5%.^32^ Participant selection followed a two-step sampling strategy. First, from the original cohort of 1066 pairs, all mother-neonate pairs with either preterm neonates or with follow-up data at 5-8 years of age were included to ensure sufficient sample size in these subgroups. Second, simple random sampling was performed among the remaining pairs.

This study was approved by the Institutional Review Board of the WHO Western Pacific Regional Office (2013.10.CHN.2.ESR), the Chinese Centre for Disease Control and Prevention (201224), the London School of Hygiene & Tropical Medicine (15698) and School of Public Health, Fudan University (IRB#2019-15-0756; #2020-11-0857; #2020-11-0857-S and #2022-02-0947). All enrolled mothers provided written informed consent for themselves and their neonates.

### Laboratory procedures

Endpoint titers of RSV pre-F IgG antibodies were obtained using an enzyme-linked immunosorbent assay (ELISA). Briefly, 96-well plates were coated with stabilized pre-F protein, then serial 4-fold diluted serum was added to the wells and incubated at 37°C for 2 hours. Horseradish peroxidase (HRP)-conjugated anti-human IgG was added, followed by addition of substrate and termination of the reaction with 0.5 M sulfuric acid. A four-parameter logistic model (4PL model) was used to fit the dilution curves for individual samples. The RSV pre-F IgG antibody titer was calculated as the final dilution at which the sample absorbance was closest to the optical density of 0.1. Additionally, to assess the agreement between RSV pre-F IgG antibodies and RSV nAbs. we randomly selected maternal-neonate pairs with at least six follow-up measurements and all preterm neonates were selected. A focus reduction neutralization test (FRNT) was used for testing RSV nAb titers. The laboratory assays have been described previously and are detailed in Supplementary Methods.^19^

### Statistical analysis

We provided descriptive statistics and estimated the GMTs among all included mother-neonate pairs. The transplacental transfer ratio of maternal antibodies was defined as the geometric mean ratios of neonate-to-mother titers (GMRs). Spearman correlation coefficients between maternal and neonatal antibody titers, as well as between maternal titers and transfer ratios were estimated, respectively. Generalized linear models (GLMs) were used to explore potential factors associated with neonatal antibody levels, as well as the transplacental transfer efficiency of maternal antibodies. Specifically, univariate analysis was first performed to explore factors potentially associated with these parameters. Then, a multivariate model was then developed to quantify the impact of each factor, given as

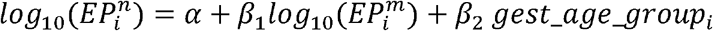

Where 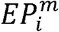 and 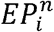 represent maternal and neonatal antibody titers for the *i*^*th*^ mother-neonate pairs, respectively, and *gest_age_group* represents the categorical variable for gestational age (0 = full-term, 1 = preterm, 2 = post-term). Accordingly, the transplacental transfer ratio (*TR*_*i*_) could be expressed as

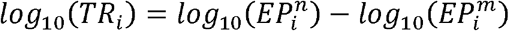

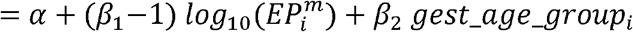

A generalized additive mixed-effects model (GAMM) accounting for individual random effects was applied to fit antibody dynamics from birth to 42 months of age. Data from the last follow-up time point (i.e. 5-8 years of age) were excluded from model fitting because the lack of follow-up between 2018 and 2021 created an 18-month gap between ages 3.5 and 5 years.

A generalized linear mixed-effects model (GLMM) was used to quantify the waning immunity of maternal antibodies, given as

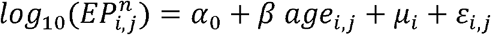

Where *age* _*i,j*_ and 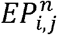 denote the neonatal age and antibody titer of the *i*^*th*^ mother-neonate pairs at the *j*^*th*^ visit, respectively, *µ*_*i*_ represents the individual random effect; and ℰ_*i,j*_ is the residual error term. *α*_0_ represents the population-level mean antibody titer at birth and *β* denotes the decay rate of maternally derived antibodies, both expressed on the *log*_10_ scale. Based on the GLMM model, the half-life (*t*_1/2_) of RSV pre-F IgG antibodies was calculated as:

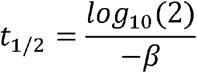

Only infants’ data within 6-month of age and without seroconversion or 2-fold titer rise was used for this analysis. Seroconversion was defined as a change from negative to positive titers between paired samples using a seropositivity threshold of 1:200.

All analyses were performed in R version 4.4.0 (R Foundation for Statistical Computing, 467 Vienna, Austria, https://www.r-project.org/). The 4PL model was performed using the drc package (version 3.0-1).^45,46^

## Contributors

H.Y. conceived, designed, and supervised the study. M.X., Q.W., X.T., J.L., and J.Y. participated in data and sample collection. M.X., Q.W., L.Y. and S.Z. performed the laboratory tests. Y.W., M.X., and Q.W. analyzed the data. Q.W., Y.W., and M.X. prepared the figures and the first draft of the manuscript. H.Y.C. made valuable revision of the manuscript for important intellectual content. All authors had full access to all the data in the study and accept responsibility for the decision to submit for publication.

## Declaration of interests

H.Y. has received research funding from Sanofi Pasteur, Shenzhen Sanofi Pasteur Biological Products Co., Ltd, Shanghai Roche Pharmaceutical Company, and SINOVAC Biotech Ltd. None of the research funding is related to this work.

H.Y.C. reports consulting with Roche, Vir, and Merck.

All other authors report no competing interests.

## Acknowledgments

H.Y. acknowledges financial support from the Key Program of the National Natural Science Foundation of China (82130093). We thank all study participants for their cooperatively participating in the study. We thank staff members of the Anhua County CDC for providing assistance with administration and data collection. We also acknowledge the valuable contributions of Dr. Barney Graham’s team at NIAID for serological assay support. We thank Dr. Anna Funk for her English language editing. The computations in this research were performed using the CFFF platform of Fudan University.

## Role of the funding source

The funder of the study had no role in the study design, data collection, data analysis, data interpretation, or writing of the report.

## Notes

### Author Declarations

This study was approved by the Institutional Review Board of the World Health Organization Western Pacific Regional Office (2013.10.CHN.2.ESR), the Chinese Centre for Disease Control and Prevention (201224), the London School of Hygiene & Tropical Medicine (15698) and School of Public Health, Fudan University (IRB#2019-15-0756; #2020-11-0857; #2020-11-0857-S and #2022-02-0947). All enrolled mothers provided written informed consent for themselves and their neonates.

