## Supplemental Information for "Transplacental transfer efficiency and longitudinal dynamics of antibodies against RSV in Chinese children from birth to 8 years: a paired mother–neonate cohort study"

**This file includes:**

**Supplementary methods**

**Table S1-S4**

**Figure S1-S4**

### Supplementary methods

#### RSV pre-F enzyme-linked immunosorbent assay (ELISA)

RSV pre-F IgG antibodies were quantified using endpoint enzyme-linked immunosorbent assay (ELISA) with minor modifications.^1^ The stabilized pre-F conformation of the F protein was purchased from Sino Biological, China. Plates were coated with RSV pre-F protein (0.4 μg/mL), and incubated overnight at 4°C. The serum specimens were heated at 56°C for 30 minutes prior to serial fourfold dilutions starting at 1:200 in diluted buffer. To minimize the impact of random experimental error, the ELISA plate layout was designed to accommodate paired maternal and serial neonatal sera on the same plate whenever possible. Samples were distributed across two serially numbered plates when a single plate's well capacity was exceeded. A serially diluted set of positive controls and negative controls and blank wells were included in each run to monitor the reproducibility of the results. An in-house positive control was prepared using a pool of remnant sera from 15 PCR-diagnosed RSV-infected inpatients aged between 2 and 4 years old from a hospital-based study. All samples were run in duplicate.

An OD value of 0.1 was established as the detection limit in the ELISA assay. The endpoint titers of IgG antibodies were determined as the final dilution where the sample's absorbance was closest to 0.1, using drc package in R (version 4.4.0) to fit a four-parameter logistic curve. The log-transformed reciprocal of the lowest dilution was used as the antibody-positive cut-off, therefore, the log_10_ value of titers less than 2.301 (log_10_200) were considered seronegative, while those with a log_10_ value of 2.301 or higher were considered seropositive. Serum samples that did not reach an absorbance of 0.1 at the initial serum dilution were assigned a titer of 1:50.

#### RSV focus reduction neutralization test (FRNT)

To assess the agreement between RSV pre-F IgG and nAb levels, we randomly selected maternal-neonate pairs with at least six follow-up measurements and all preterm neonates. A Focus reduction neutralization test (FRNT) was conducted. Briefly, this involved mixing serial dilutions of heat-inactivated sera with RSV A strain (A2, ATCC) separately for 120 minutes at 4℃ and then transferring the mixture onto a Vero (CCL-81, ATCC) epithelial cell monolayer in a 96-well tissue culture plate. After incubating for 1.5 hours at 37℃ in 5% CO2, the inoculum was removed, and each well was overlaid with a final concentration of 2% carboxymethylcellulose (low viscosity, Sigma-Aldrich, St. Louis, MO). After 48 hours, viral foci were detected using a goat polyclonal antibody that binds RSV fusion glycoprotein (AB1128, Millipore Inc., US) followed by an HRP-conjugated donkey anti-goat IgG secondary antibody (A0181, Beyotime, China). Plates were then scanned and viral foci were counted using the image analyzer, EliSpot Reader (AID Diagnostika GmbH, Germany). All FRNT experiments were performed in an approved Biosafety Level 2 setting.

The RSV neutralizing antibody titer was expressed as the dilution of serum that resulted in 50% foci reduction (FRND50), and was calculated using a previously developed Bayesian hierarchical framework.^2^ We included the first International Standard for Antiserum to RSV in each batch of FRNT. By multiplying our neutralizing antibody by the conversion factor of 2.37, we were able to approximate WHO International Units (IU/mL) for RSV A.

**Potential impact of COVID-19 pandemic**

To evaluate the potential impact of COVID-19 pandemic on population-level RSV immunity, we combined data from this study with subsets of data collected in two independent studies conducted in Anhua county (the same study site as the present work). One of the studies was a community-based serological cohort established in 2013, which enrolled local children aged 1-9 years and conducted seven follow-ups in Feb-Mar 2014, Aug-Oct 2014, Mar 2015, Aug-Oct 2015, Mar 2016, Aug-Nov 2016, and Jul-Oct 2021. Serum samples were collected at enrollment and at each follow-up visit. The other study was a community-based cross-sectional sero-epidemiological survey conducted from Jul-Nov 2021, and included participants aged 4 months to older than 89 years. Both studies have been previously published^1,3,4^. From these two studies, we randomly selected serum samples from children under 8 years of age (sample sizes for each age group are shown in Fig. S3B) We measured RSV pre-F IgG antibody titers and compared levels before and after the pandemic, with the results shown in Fig. S3A.

#### Table S1. Characteristics between selected and non-selected participants from the original cohort

| **Characteristics** | **Total** | **Selected participants** | **Non-selected participants** | ***P* value** |
| --- | --- | --- | --- | --- |
| Mothers | N = 1054 | N = 687 | N = 367 |  |
| Age at delivery (years) |  |  |  | 0.577 |
| Median age (IQR) | 25.0 (23.0-29.0) | 25.0 (23.0-29.0) | 25.0 (23.0-29.0) |  |
| 16-19 | 40 (3.8) | 25 (3.6) | 15 (4.1) |  |
| 20-24 | 393 (37.4) | 266 (38.7) | 127 (34.8) |  |
| 25-29 | 375 (35.6) | 240 (34.9) | 135 (37.0) |  |
| 30-34 | 170 (16.2) | 107 (15.6) | 63 (17.3) |  |
| 35-44 | 73 (6.9) | 49 (7.1) | 24 (6.6) |  |
| 45-49 | 1 (0.1) | 0 (0.0) | 1 (0.3) |  |
| Gravidity |  |  |  | 0.773 |
| 1 | 321 (30.5) | 205 (29.8) | 116 (31.6) |  |
| 2 | 431 (40.9) | 286 (41.6) | 145 (39.5) |  |
| ≥ 3 | 296 (28.1) | 193 (28.1) | 103 (28.1) |  |
| Missing | 6 (0.6) | 3 (0.4) | 3 (0.8) |  |
| Parity |  |  |  | 0.116 |
| 1 | 498 (47.2) | 313 (45.6) | 185 (50.4) |  |
| 2 | 518 (49.1) | 345 (50.2) | 173 (47.1) |  |
| ≥ 3 | 32 (3.0) | 26 (3.8) | 6 (1.6) |  |
| Missing | 6 (0.6) | 3 (0.4) | 3 (0.8) |  |
| Mode of delivery |  |  |  | 0.308 |
| Vaginal delivery | 666 (63.2) | 426 (62.0) | 240 (65.4) |  |
| Caesarean section | 388 (36.8) | 261 (38.0) | 127 (34.6) |  |
| Neonates | N = 1066 | N = 695 | N = 371 |  |
| Sex |  |  |  |  |
| Male | 576 (54.0) | 367 (52.8) | 209 (56.3) | 0.300 |
| Female | 490 (46.0) | 328 (47.2) | 162 (43.7) |  |
| Gestational age at birth, weeks | |  |  | <0.001 |
| Median (IQR) | 40.0 (39.1, 40.7) | 40.0 (39.0, 40.7) | 40.0 (39.3, 40.8) |  |
| Preterm birth (<37)* | 27 (2.5) | 27 (3.9) | 0 (0.0) |  |
| Full-term birth (37-42) | 998 (93.6) | 644 (92.7) | 354 (95.4) |  |
| Post-term birth (>42) | 41 (3.8) | 24 (3.5) | 17 (4.6) |  |
| Birthweight, kilograms |  |  |  | 0.231 |
| Median (IQR,) | 3.3 (3.0, 3.6) | 3.3 (3.0, 3.6) | 3.4 (3.1, 3.6) |  |
| < 2.5 | 27 (2.5) | 19 (2.7) | 8 (2.2) |  |
| 2.5 to < 4 | 963 (90.3) | 633 (91.1) | 330 (88.9) |  |
| ≥ 4 | 76 (7.1) | 43 (6.2) | 33 (8.9) |  |
| Having twin siblings |  |  |  | 0.350 |
| Yes | 25 (2.3) ^†^ | 6 (1.4) | 19 (2.7) |  |
| No | 1041 (97.7) | 676 (97.3) | 365 (98.4) |  |

Data are shown as n (%), unless otherwise specified. The percentages might not total 100% because of rounding. ^†^There are 12 pairs of twins, and one neonate with a twin sibling that did not participate in this cohort study. *All preterm births were included in this study.

#### Table S2. Univariate analysis of factors associated with RSV pre-F IgG antibody levels in neonates at birth

| **Characteristics** | **β (95% CI)** | **Fold changes**  **(95% CI)** | ***P* value** |
| --- | --- | --- | --- |
| Maternal antibody titers | 0.81 (0.77, 0.84) | 5.46 (4.89, 5.92) | <0.001 |
| Age at delivery (years) |  |  |  |
| 16-19 | Reference | Reference |  |
| 20-24 | 0.06 (-0.04, 0.17) | 0.15 (-0.09, 0.48) | 0.243 |
| 25-29 | 0.05 (-0.05, 0.16) | 0.12 (-0.11, 0.45) | 0.323 |
| 30-34 | 0.03 (-0.08, 0.15) | 0.07 (-0.17, 0.41) | 0.573 |
| 35-44 | 0.06 (-0.06, 0.19) | 0.15 (-0.13, 0.55) | 0.327 |
| Gravidity |  |  |  |
| 1 | Reference | Reference |  |
| 2 | -0.01 (-0.06, 0.04) | -0.02 (-0.13, 0.10) | 0.716 |
| ≥ 3 | 0.02 (-0.04, 0.07) | 0.05 (-0.09, 0.17) | 0.554 |
| Parity |  |  |  |
| 1 | Reference | Reference |  |
| 2 | -0.03 (-0.06, 0.01) | -0.07 (-0.13, 0.02) | 0.210 |
| ≥ 3 | -0.02 (-0.12, 0.09) | -0.05 (-0.24, 0.23) | 0.733 |
| Mode of delivery |  |  |  |
| Vaginal delivery | Reference | Reference |  |
| Caesarean section | -0.02 (-0.05, 0.02) | -0.05 (-0.11, 0.05) | 0.456 |
| Sex |  |  |  |
| Male | Reference | Reference |  |
| Female | 0.03 (-0.01, 0.07) | 0.07 (-0.02, 0.17) | 0.146 |
| Gestational age at birth, weeks | |  |  |
| Continuous gestational age | 0.00 (-0.01, 0.02) | 0.00 (-0.02, 0.05) | 0.430 |
| Preterm birth (<37) | -0.01 (-0.11, 0.09) | -0.02 (-0.22, 0.23) | 0.798 |
| Full-term birth (37-42) | Reference | Reference |  |
| Post-term birth (> 42) | 0.03 (-0.08, 0.13) | 0.07 (-0.17, 0.35) | 0.609 |
| Birthweight, kilograms |  |  |  |
| < 2.5 | -0.06 (-0.18, 0.06) | -0.13 (-0.34, 0.15) | 0.318 |
| 2.5 to < 4 | Reference | Reference |  |
| ≥ 4 | 0.05 (-0.03, 0.13) | 0.12 (-0.07, 0.35) | 0.201 |

We quantified RSV pre-F IgG antibodies using log_10_ endpoint titers. *β* > 0 indicates that the factor was associated with an increase in RSV pre-F IgG antibodies by *10^β^*-1-fold; *β* < 0 indicates that the factor was associated with a decrease in RSV pre-F IgG antibodies by 1-*10^β^* fold.

#### Table S3. Univariate analysis of factors associated with the transfer ratios of maternal RSV pre-F IgG antibodies

| **Characteristics** | **β (95% CI)** | **Fold changes**  **(95% CI)** | **P value** |
| --- | --- | --- | --- |
| Maternal antibody titers | -0.19 (-0.23, -0.16) | -0.35 (-0.41, -0.31) | <0.001 |
| Age at delivery (years) |  |  |  |
| 16-19 | Reference | Reference |  |
| 20-24 | -0.03 (-0.08, 0.03) | -0.07 (-0.17, 0.07) | 0.400 |
| 25-29 | -0.02 (-0.08, 0.04) | -0.05 (-0.17, 0.10) | 0.522 |
| 30-34 | -0.03 (-0.09, 0.03) | -0.07 (-0.19, 0.07) | 0.385 |
| 35-44 | -0.02 (-0.09, 0.04) | -0.05 (-0.19, 0.10) | 0.484 |
| Gravidity |  |  |  |
| 1 | Reference | Reference |  |
| 2 | 0.00 (-0.03, 0.02) | 0.00 (-0.07, 0.05) | 0.832 |
| ≥ 3 | 0.00 (-0.03, 0.03) | 0.00 (-0.07, 0.07) | 0.871 |
| Parity |  |  |  |
| 1 | Reference | Reference |  |
| 2 | -0.02 (-0.04, 0.00) | -0.05 (-0.09, 0.00) | 0.104 |
| ≥ 3 | -0.02 (-0.08, 0.04) | -0.05 (-0.17, 0.10) | 0.515 |
| Mode of delivery |  |  |  |
| Vaginal delivery | Reference | Reference |  |
| Caesarean section | 0.00 (-0.02, 0.03) | 0.00 (-0.05, 0.07) | 0.716 |
| Sex |  |  |  |
| Male | Reference | Reference |  |
| Female | 0.00 (-0.02, 0.03) | 0 (-0.05, 0.07) | 0.680 |
| Gestational age at birth, weeks | |  |  |
| Continuous gestational age | 0.01 (0.00, 0.02) | 0.02 (0.00, 0.05) | 0.001 |
| Preterm birth (<37) | -0.09 (-0.15, -0.04) | -0.19 (-0.29, -0.09) | 0.001 |
| Full-term birth (37-42) | Reference | Reference |  |
| Post-term birth (> 42) | 0.00 (-0.06, 0.06) | 0.00 (-0.13, 0.15) | 0.957 |
| Birthweight, kilograms |  |  |  |
| < 2.5 | -0.04 (-0.10, 0.03) | -0.09 (-0.21, 0.07) | 0.283 |
| 2.5 to < 4 | Reference | Reference |  |
| ≥ 4 | -0.01 (-0.05, 0.03) | -0.02 (-0.11, 0.07) | 0.657 |

We quantified RSV pre-F IgG antibodies using log_10_ endpoint titers. *β* > 0 indicates that the factor was associated with an increase in transplacental transfer ratios of maternal RSV pre-F IgG antibodies by 10*^β^*-1-fold; *β* < 0 indicates that the factor was associated with a decrease in transplacental transfer ratios of maternal RSV pre-F IgG antibodies by 1-10*^β^* fold.

#### Table S4. Characteristics of study participants selected for neutralization tests.

| **Characteristics** |  |
| --- | --- |
| **Mothers** | N = 163 |
| Age at delivery, years |  |
| Median (interquartile range, IQR) | 25.0 (23.0, 29.5) |
| Gravidity |  |
| 1 | 49 (30.1) |
| 2 | 71 (43.6) |
| ≥3 | 42 (25.8) |
| Missing | 1 (0.6) |
| Parity |  |
| 1 | 71 (43.6) |
| 2 | 83 (50.9) |
| ≥3 | 8 (4.9) |
| Missing | 1 (0.6) |
| **Neonates** | N = 167 |
| Sex |  |
| Male | 90 (53.9) |
| Female | 77 (46.1) |
| Gestational age at birth, weeks |  |
| Median (IQR) | 39.6 (38.5, 40.6) |
| Preterm birth (< 37) | 27 (16.2) |
| Full-term birth (37 - 42) | 138 (82.6) |
| Post-term birth (> 42) | 2 (1.2) |
| Birthweight, kilograms |  |
| Median (IQR) | 3.3 (2.9, 3.5) |
| < 2.5 | 9 (5.4) |
| 2.5 to < 4 | 158 (94.6) |
| ≥ 4 | 0 (0) |
| Having twin siblings |  |
| Yes | 9 (5.4) |
| No | 158 (94.6) |

**Note:** Data are shown as n (%), unless otherwise specified. Percentages might not total 100% because of rounding. There are 4 pairs of twins, and 1 neonate who had a twin sibling that did not participate in the study.

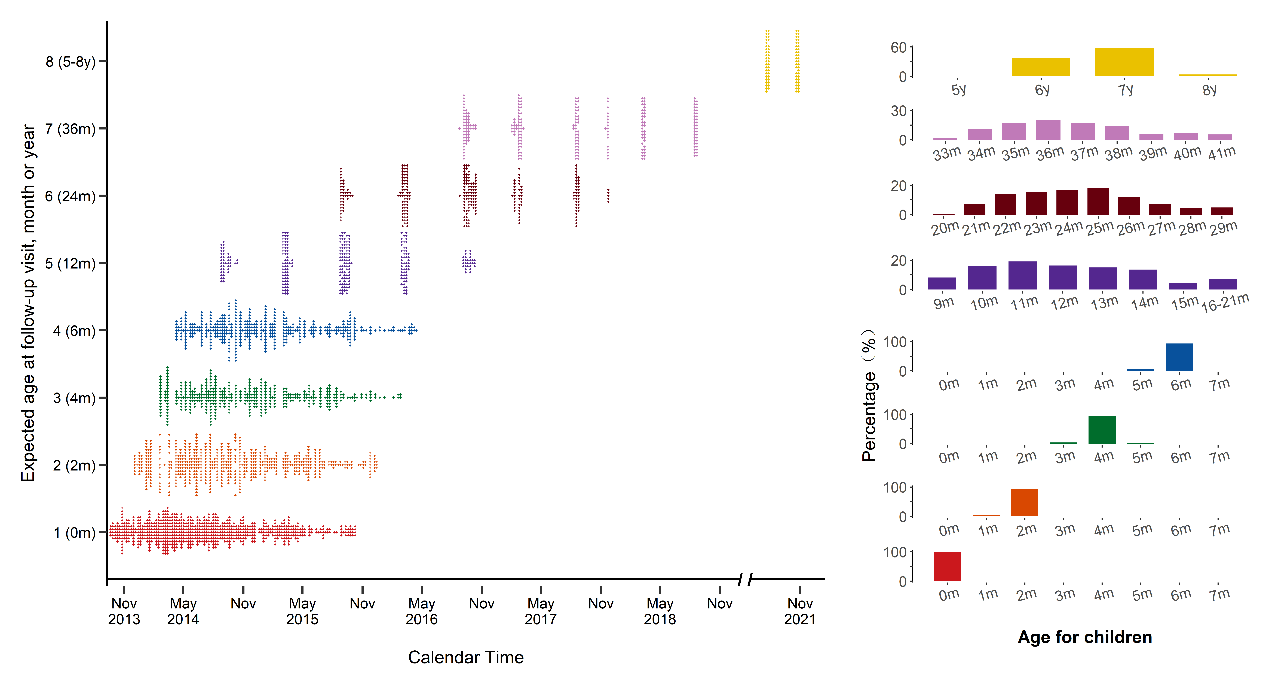
**Figure S1 Distribution of b**lood sampling and age in each follow-up visit for cohort participants

Panel left: each point is the time of blood sampling for each individual; Panel right: the numbers below the x-axis refer to the age (months or years) of participants at the follow-up time-point; each bar indicates the percentage of children in each age group.

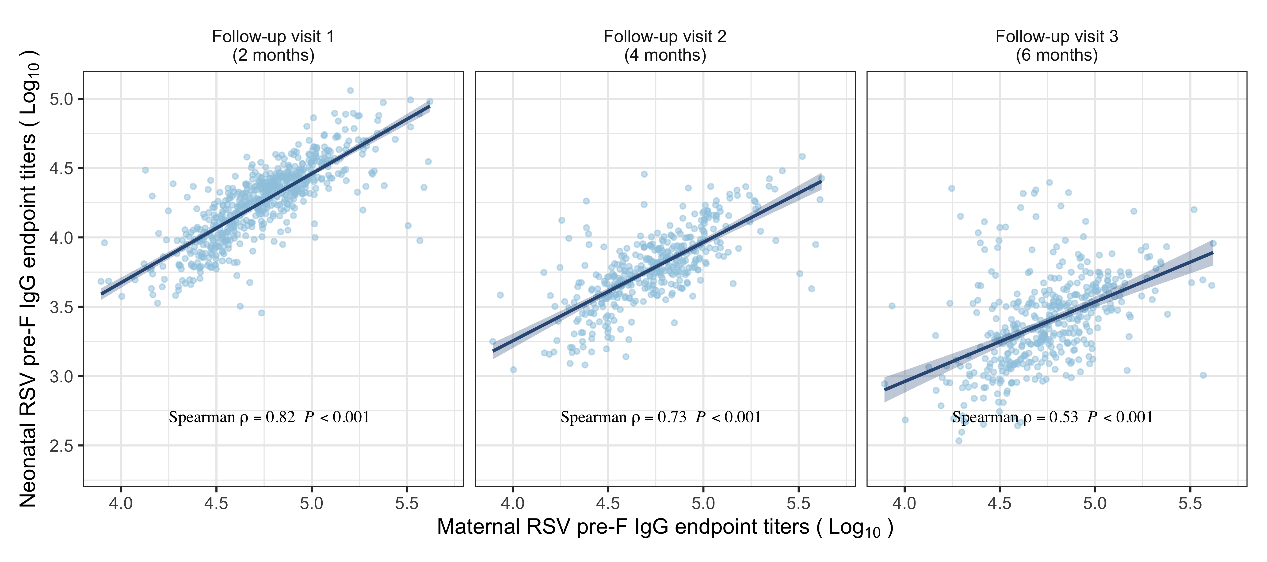
**Figure S2 Correlation between maternal antibody titers and neonatal antibody titers at around 2, 4, and 6 months of age**

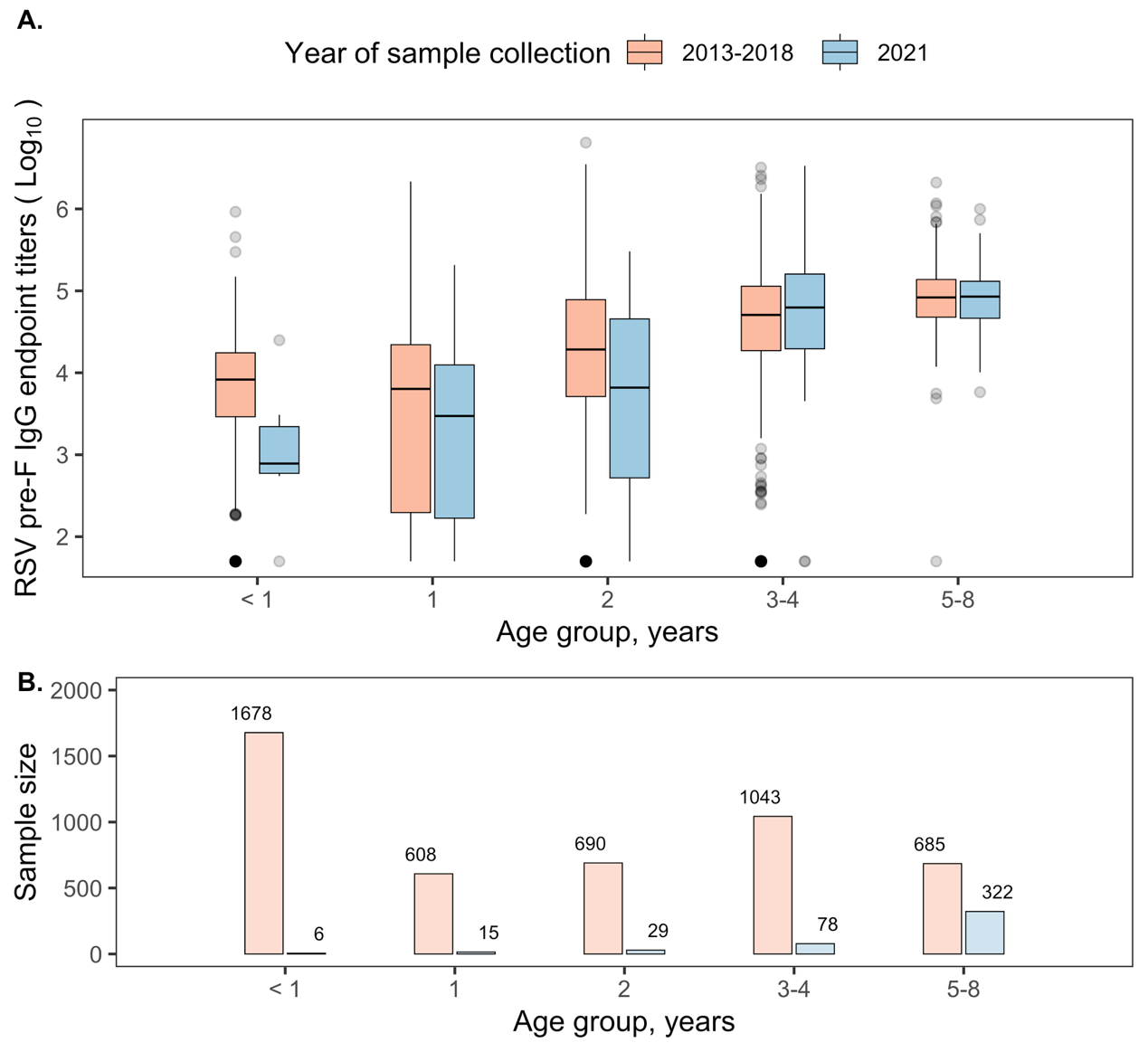

**Figure S3 RSV pre-F IgG antibody levels in community-based serum samples collected before and during the COVID-19 pandemic** (A) Age-specific GMTs. (B) Sample size in each age group.

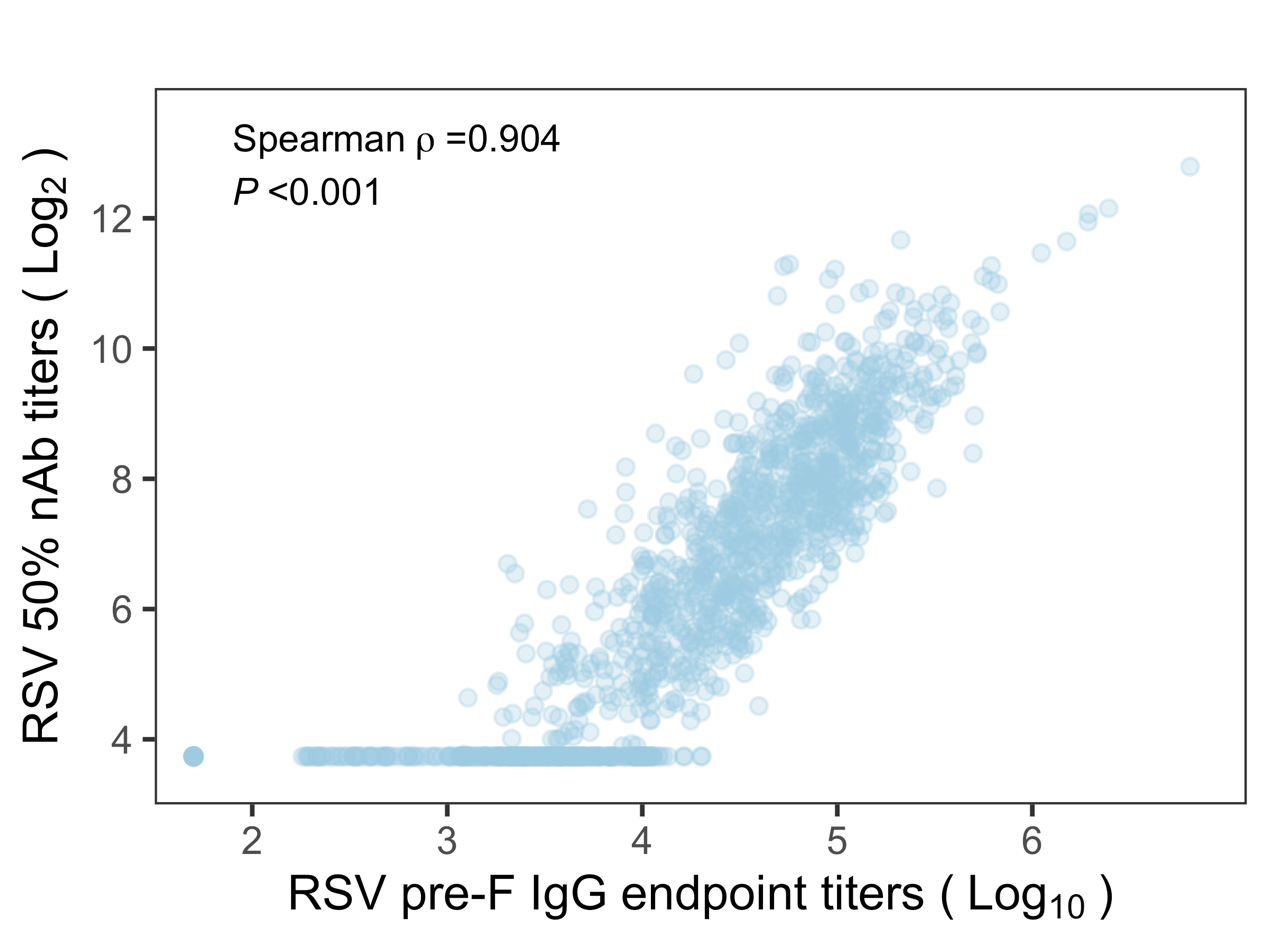

**Figure S4 Correlation between RSV pre-F IgG antibody titers and neutralization antibody titers**
